# Compositional Cyber-Physical Epidemiology of COVID-19

**DOI:** 10.1101/2020.04.26.20081125

**Authors:** Jin Woo Ro, Nathan Allen, Weiwei Ai, Debi Prasad, Partha S. Roop

## Abstract

COVID-19 pandemic has posed significant challenges globally. Countries have adopted different strategies with varying degrees of success. Epidemiologists are studying the impact of government actions using scenario analysis. However, the interactions between the government policy and the disease dynamics are not formally captured.

We, for the first time, formally study the interaction between the disease dynamics, which is modelled as a physical process, and the government policy, which is modelled as the adjoining controller. Our approach enables compositionality, where either the plant or the controller could be replaced by an alternative model. Our work is inspired by the engineering approach for the design of Cyber-Physical Systems (CPSs). Consequently, we term the new framework Compositional Cyber-Physical Epidemiology (CCPE). We created different classes of controllers and applied these to control the disease in New Zealand and Italy. Our controllers closely follow government decisions based on their published data. We not only reproduce the pandemic progression faithfully in New Zealand and Italy but also show the tradeoffs produced by differing control actions.

---

The ongoing Coronavirus Disease 2019 (COVID-19) presents an unprecedented global crisis with over 2,718,155 infections and 190,636 deaths as of 24th April 2020. There are now widespread calls for new techniques for intervention, including methods of rapid testing even at the home [1]. While Epidemiologists are studying the dynamics of the diseases using computational models, governments are trying to “flatten the curve” [2] to reduce the health impacts. This is achieved through Nonpharmaceutical Interventions (NPIs), such as lockdowns and social distancing methods.

The Majority of the research focus in epidemiology has been on mathematical modelling of the disease dynamics. Many governments, like the New Zealand government, have also worked closely with the scientific community [3] to arrive at critical decisions. But how can we ascertain which model is better, and in which settings [4]? There exists no clear methodology to formally *capture and classify criteria-based actions of the government* [5] as mathematical models. Also, *given the wide variability of government actions globally, how can we formally assess them while studying their impact*? This paper tries to provide answers to these questions for the first time, by focusing on the compositional modelling of government actions alongside an epidemiological model of the disease.

While at the policy level there has been minimal engineering thinking to provide solutions, it is evident that the pandemic and its control bears many similarities with the well known engineering domain of Cyber-Physical Systems (CPSs) [6], [7]. In a CPS, a physical process such as the electrical conduction of the human heart (known as the *Plant*) is controlled by an adjoining device such as a pacemaker, also known as a *Controller* [8]. This closed-loop system mimics the behaviour of a piece-wise continuous phenomena, where the plant’s dynamics is modelled using a set of Ordinary Differential Equations (ODEs). The plant makes discrete mode switches based on the actions of a discrete controller.

In the setting of COVID-19, we may view the plant as the dynamics of disease progression, already modelled faithfully using several epidemiological models [9], [10]. The adjoining controller is a state machine that can induce mode switches in the plant. Such a closed-loop system may be depicted as shown in Figure 1a and we term this approach Compositional Cyber-Physical Epidemiology (CCPE). Here, the plant provides the state of the pandemic encapsulated as a vector of variables *X*(*t*), while the controller affects the state of the plant by trying to alter the value of the reproduction number *R*_0_, which represents the average number of new infections for each infectious person.

**Fig. 1:**
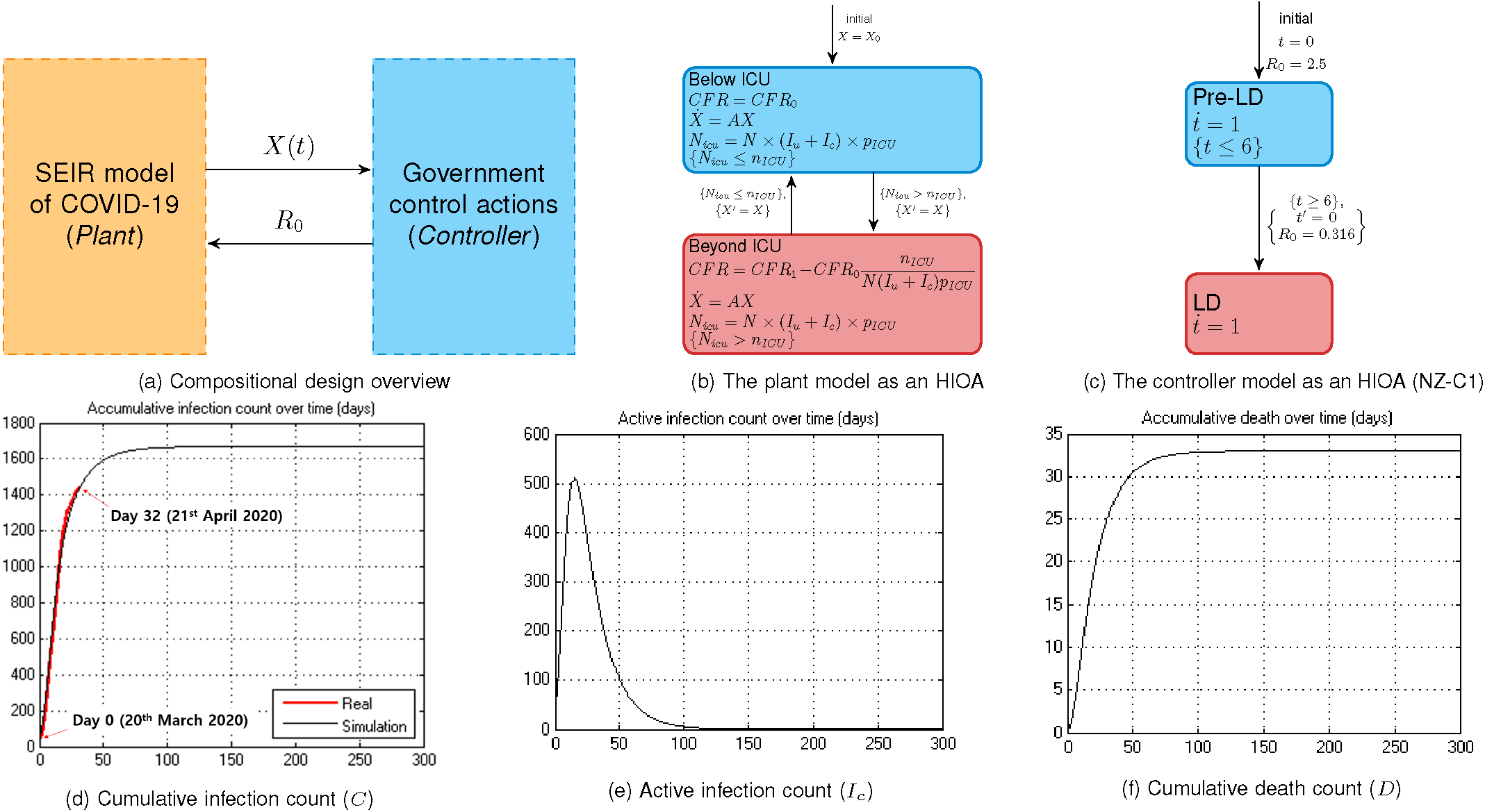
The proposed compositional design of Compositional Cyber-Physical Epidemiology and simulation results. For (d), (e), and (f), day 0 corresponds to 20th March 2020. The first 32 days of (d) are compared with the available New Zealand data.

There is recent evidence that such engineering thinking may have relevance for COVID-19. The Institute of Electrical and Electronics Engineers (IEEE) published an article citing the benefits of the application of such feedback control theory [11], which is evidence of concurrent thinking along our lines. However, their work is primarily based on studying the impact of fictitious controllers over a simple disease model, without considering the actual data from the current pandemic.

## A. Plant and Controller Dynamics

This paper advocates that a compositional design approach is needed to include the NPI techniques with the existing epidemiological models. Such an approach, which we term *CCPE*, would allow for the creation of more realistic models which can answer more questions than any existing individual model [4]. This can be used in the decision making of a government, with the goal of both minimising the death toll while reducing the economic impact of any restrictions.

We use a Susceptible, Exposed, Infected, Removed (SEIR) model [9], [10] for the illustration of our methodology, while stressing that the developed methodology is amenable to any other dynamical model based on ODEs. The SEIR model incorporates coupled ODEs, and has been utilised previously in the context of COVID-19 [12]. These ODEs capture the progression of a disease through the population, as people become infected, progress through their infection, and infect others. SEIR models include variables which represent the population during an epidemic which can be in a range of states: susceptible (*S*), exposed (*E*), pre-symptomatic (*P*), infectious (*I*), recovered (*R*) and deaths (*D*). The infected and recovered cases are further categorized into untested (*I_u_*, *R_u_*) and confirmed cases (*I_c_*, *R_c_*) to enable control mechanisms which are specific to confirmed cases. The key parameter determining if a virus can cause an epidemic is the reproduction number *R*_0_ and depends on both the transmissibility of the virus and social distancing. For *R*_0_ > 1 the virus will spread until herd immunity has been established, while for *R*_0_ < 1 the transmission will progressively decay until the virus is eradicated [10]. In addition to *R*_0_, further parameters are used for capturing aspects such as the fatality rate and testing rate.

Government interventions can be used to modify each of these parameters such as the use of NPIs to reduce the reproduction number, or increased testing to isolate more confirmed cases. These responses vary between countries and typically vary over time depending on the local situation [13], [14]. For example, New Zealand has implemented an alert system for COVID-19 [15] which comprises four levels of increasingly strict interventions. In this case, the four levels can be modelled as a discrete controller [7] which can interact with the continuous SEIR model as a type of CPS.

## B. Formal Modelling

The use of formal modelling for biological processes has been advocated by Fisher and Henzinger [16], which makes a distinction between computational models and *executable models*. More recently, Bioengineers have adopted an executable model called Hybrid Input-Output Automata (HIOAs) [8], [17] for developing abstract models. These abstractions are used to achieve behaviour from cellular [18] to organ levels [19], [20]. These abstract models are also “executable” in the sense that hardware and software implementations may be derived from them so that they work as virtual organs [21], [19], [22].

An HIOA captures both the continuous (i.e. the population model) and discrete (i.e. the government controller) dynamics through the use of an automata with included ODEs. The conversion of the SEIR model into an HIOA results in the formal model of Figure 1b, where the two locations capture whether the Intensive Care Unit (ICU) capacity has been exceeded. The formal nature of these models means that they can be used in simulation and code generation frameworks with relative ease [23], [24]. We have used the recently developed Hybrid Automata Modelling Language (HAML) [21], which allows for the specification of a complex network of HIOAs.

To illustrate our methodology we have selected New Zealand and Italy, who have adopted contrasting approaches in the disease management. We show that in the case of the four level New Zealand control, we are able to make decisions around the optimal criteria for switching between the control modes to minimise the impact of the virus. Our methodology is generic enough and has the potential to be adopted to other alternative settings.

## Results

### A simple controller in the New Zealand context

The proposed CCPE approach is first demonstrated using the New Zealand COVID-19 context using a simple controller we term `NZ-C1`. The `NZ-C1` control strategy is to initiate a strong lockdown measure, which is introduced early and is not lifted until the new infections approach zero. We use the dataset, which contains the number of cases (both confirmed and probable), recovered, and deaths for every day from 20th March 2020 to 21st April 2020 (overall 33 days including the starting day). Also, on 26th March 2020 (six days after the first date in our data), the New Zealand government initiated their *level four lockdown measures*, which are scheduled to be in place until 27th April 2020. Then, *level three* starts from 28th April 2020.

First, we examine the accuracy of our CCPE approach by comparing it to the New Zealand data as a means to increase confidence in our predicted future disease dynamics, as per Figure 1. In our framework, we propose the modelling of both the plant and controller as HIOA [7]. We create a simple controller (Figure 1c), which transitions into a lockdown mode (`LD`) six days after the start. The controller modes are depicted as two different states of the system, namely `Pre-LD` and `LD` respectively. Within every mode, we encapsulate a condition that determines the maximum time control can reside in a given mode, which is known as the *invariant*. In the `LD` mode, however, no such invariant is specified. In this case, the invariant is by default true and hence control can remain in this location forever. In contrast, control can remain in the `Pre-LD` mode, when the current time *t* is less than 6 days. The rate of change of time is modelled as an ODE *ṫ* = 1 within both modes.

Transitions between modes happen when some conditions are satisfied. For example, the transition from `Pre-LD` to `LD` happens when the current value of *t* becomes 6. When a transition triggers, some variables are updated. For example, when this transition triggers, the value of time *t* is reset (by the reset action, which is denoted *tʹ* = 0). Also the value of *R*_0_ is set to 0.316.

For this model, we use the previously described values for *R*_0_ of 2.5 and 0.316 for pre-lockdown and lockdown respectively. Figures 1d through 1f show the results of this simulation for three main metrics. Day 32 in the graph corresponds to the last day observed in the New Zealand data (21st April 2020). On this date, the simulated model predicts 1447 confirmed cases, while in reality there were 1445, an error of only 2 cases. Overall, the **correlation coefficient is 0.997321**.

With this simple controller which remains in lockdown indefinitely (i.e. until a vaccine arrives), the cumulative infection count converges to 1670. Furthermore, in Figure 1e, we can observe that the active infections are almost zero, meaning that the disease has been eradicated, on day 120 (4 months). Finally, the total number of deaths in this scenario is expected to be 33.

### CCPE model of the New Zealand Government control strategy

Next, we investigate the disease dynamics in New Zealand over a longer period of time (600 days) with a more complex model which closely follows the government’s strategy of four different alert levels (Figure 2). The previous controller (Figure 1c) is extended by incorporating a control policy that reflects these alert levels. This new controller is called `NZ-C2` and is shown in Figure 2a.

**Fig. 2:**
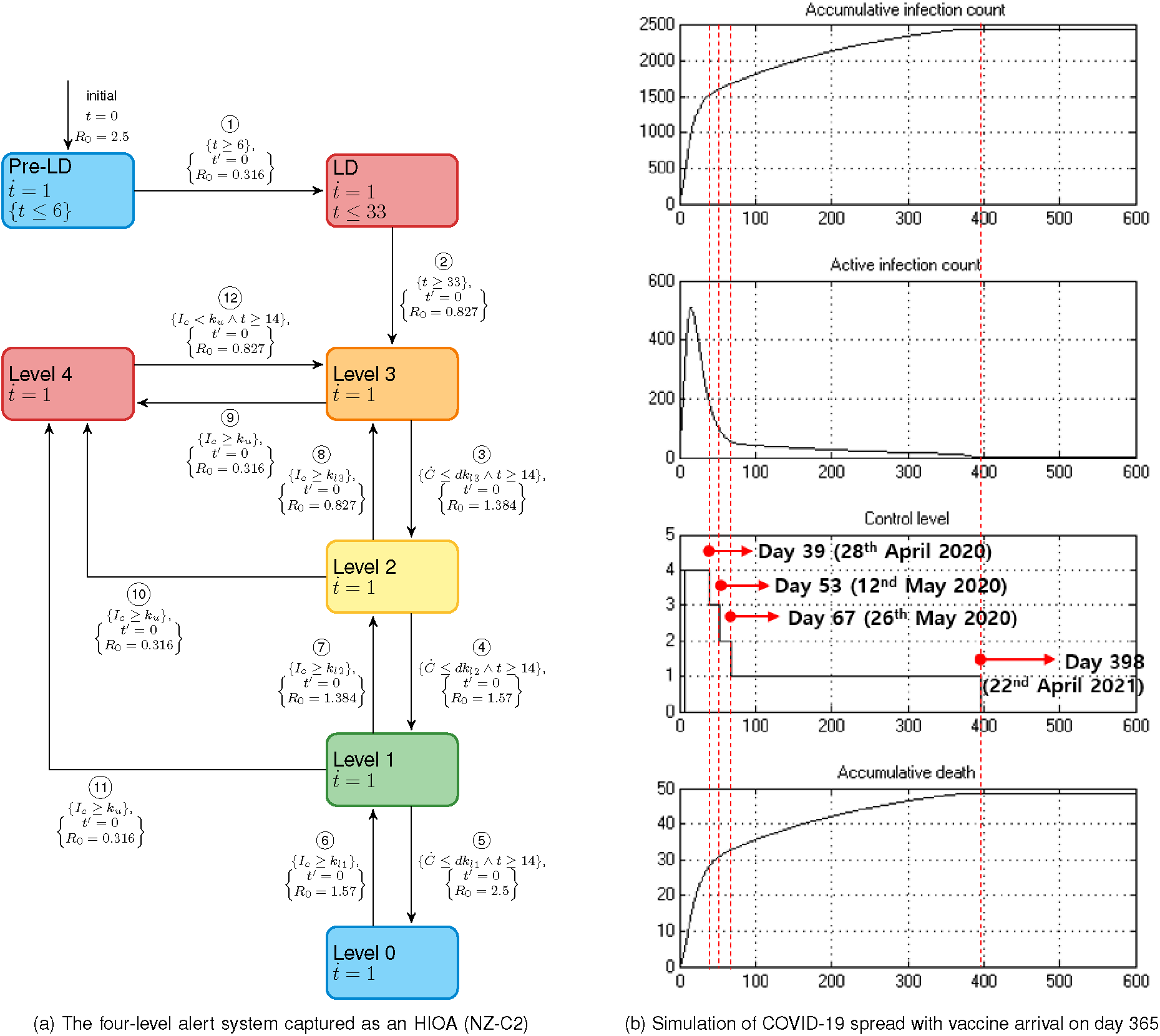
The controller and simulation results corresponding to the New Zealand system for fighting COVID-19

`NZ-C2`, in contrast to `NZ-C1`, tries to set the alert level in order to determine an appropriate reproduction number *R*_0_ for the current situation. We have based our work on the reports released by the New Zealand government and the analysis of *R*_0_ values and associated alert levels [5].

For New Zealand, Table II presented in the *Methods* section, lists major interventions and their associated relative reproduction number changes indicating how they increase/decrease the *R*_0_. According to [25], the initial value of *R*_0_ is 2.5 without any control, which corresponds to alert level 0 in our model. In summary, the *R*_0_ values for alert levels 4 through 1 are 0.316, 0.827, 1.384, 1.570 respectively. The maximum value of *R*_0_ is 2.5, which corresponds to level 0.

The controller HIOA which captures the transitions between these levels is shown in Figure 2a. Here, the conditions for increasing the alert level are based on the current number of infected cases (*I_c_*). For example, from level two if *I_c_* ≥ *k_l_*_3_ then the alert level immediately rises to three. On the other hand, the alert level can go down if the increasing rate of new cases per day (*Ċ*) is less than a certain amount. For example, from level three if *Ċ* ≤ *dk_l_*_3_ then the alert level decreases to level two. In addition, to avoid frequent oscillations between levels, a minimum duration within a level before being able to drop down to a lower level is added and is set to be 14 days.

The simulation results for this controller are shown in Figure 2b, where we also include the presence of a vaccine from day 365. In contrast to the scenario of continuing the lockdown based on the previous controller `NZ-C1`, we observe gradual step downs in the control level. Although there will be 49 deaths, 16 more than the previous lockdown scenario, the four-level approach allows for society to begin its return to normalcy from day 39 in order to minimise economic damage relative to the controller `NZ-C1`.

### Modelling Italy’s control strategy

The CCPE approach can be adapted to the intervention techniques of other countries. For example in Italy, the government does not have an explicitly outlined intervention system, instead the control actions are progressively released as they are needed. We use the published *stringency index* [13] for Italy across time in order to create an approximation of their control strategy in our framework. For example, on 23rd February 2020, the stringency index was listed as 66.67, while subsequent measures increased this to 71.43, 90.48, and finally 95.24 [13]. We create an approximate discrete controller for this approach (Figure 3a), where the phases correspond to a degrees of stringency mentioned earlier. Note that the date of first observation point in the Italy data is 23rd February 2020.

**Fig. 3:**
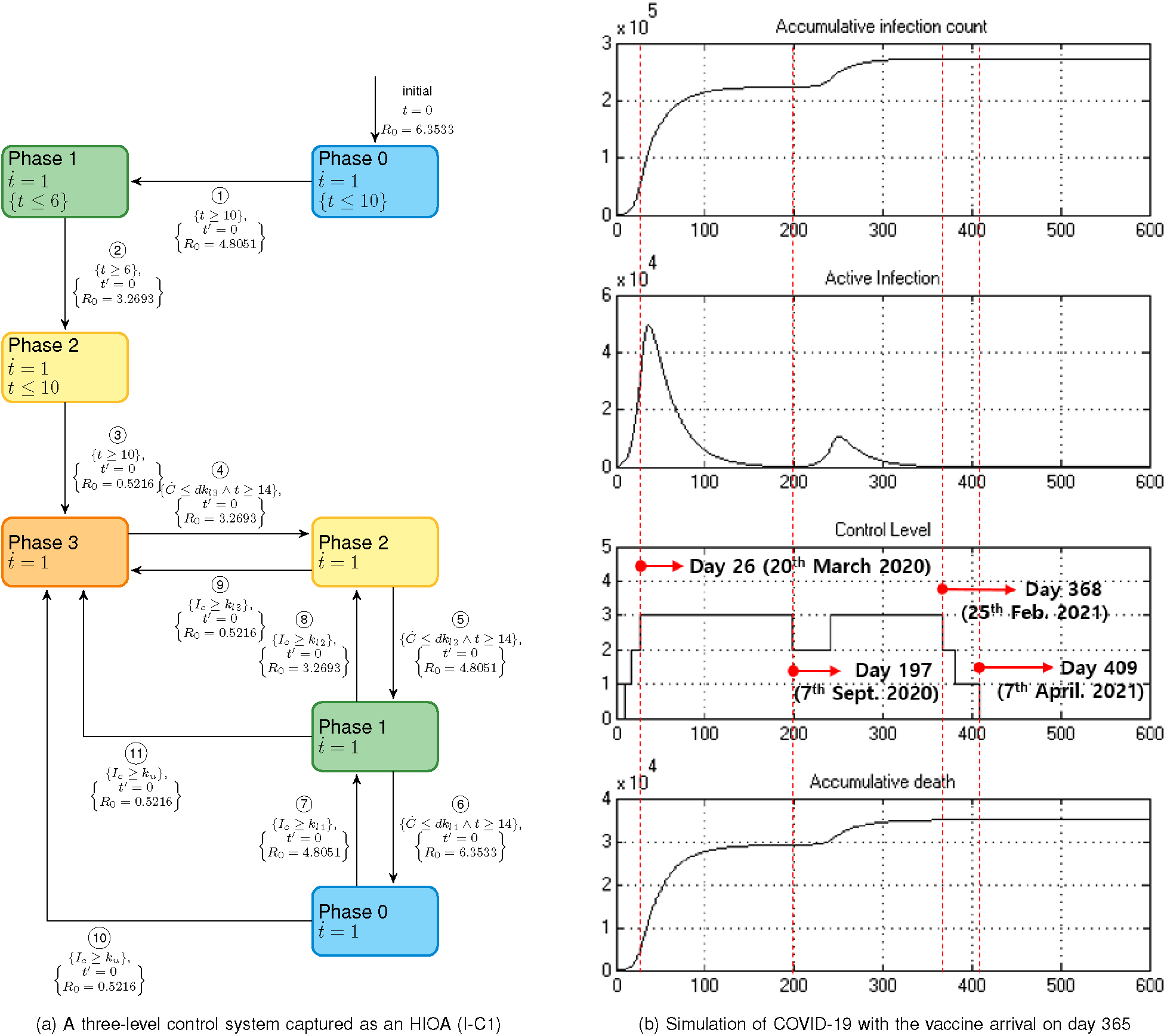
The controller and simulation results corresponding to the Italy system for fighting COVID-19

The control flow of the Italy model called `I-C1` in Figure 3a is as follow. From the initial state `Phase` 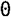, transition ① leads to `Phase 1`. This transition is triggered based on time, according to the historical actions of Italy government. For instance, Italy was in `Phase` 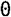 on 23rd February 2020, and moved to `Phase 1` by closing the schools and universities on 4th March 2020. Similarly, transitions ② and ③ are triggered based on the time when historical actions were imposed. Countrywide lockdown was issued on 10th March 2020 and the nation entered `Phase 2`. On 20th March 2020, the government further tightened the control by reducing the public transportation and initiated `Phase 3`. For Italy, according to our estimation based on [13], the reproduction number *R*_0_ is 6.3533 in `Phase` 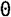, 4.8051 in `Phase 1`, 3.2693 in `Phase 2`, and 0.5216 in `Phase 3`.

From `Phase 3` in Figure 3a, we apply the same control strategy presented in Figure 2a. That is, the control level can decrease based on *Ċ* and time remained in a level, or the control level can increase based on *I_c_*. Precisely, the same level changing conditions are used for Italy. In this way, we can examine the performance of the same controller in different countries. We set 10, 5, and 0.01 for *dk_l_*_3_, *dk_l_*_2_, and *dk_l_*_1_, respectively. Also, *k_l_*_3_, *k_l_*_2_, and *k_l_*_1_ are 6046, 3023, and 605, respectively. Additionally, the constraint to level four (*k_u_*) is equal to the hospital capacity of approximately 483,694 [12].

The simulation results for the Italy model are shown in Figure 3b. First, we can observe that the control strictness rises to phase three as per the existing data, and remains there until day 199. When control goes down to phase two, the active infection count (*I_c_*) starts to increase again, causing a second wave of infections and necessitating the return to phase three. This likely indicates that there is a need for an additional phase between three and two for Italy, which is able to contain the disease without being as strict as phase three. Overall, the simulation predicts that approximately 35,000 deaths and 273,000 confirmed cases are expected in Italy.

### Modelling other controllers

We can examine various “what if” scenarios of COVID-19 in New Zealand, as a result of varying intervention techniques. A simple control policy in previous work has consisted of only two levels, essentially a full lockdown and no control [12]. Precisely, a complete lockdown (level four) is triggered if the currently active infection count exceeds the hospital ICU capacity ({*I_c_* ≥ *k_u_*}), while in times where the currently active infection count is less than the half of the hospital capacity ({*I_c_* ≤ *k_u_*/2}), the lockdown is removed (level zero), as shown in Figure 4a.

**Fig. 4:**
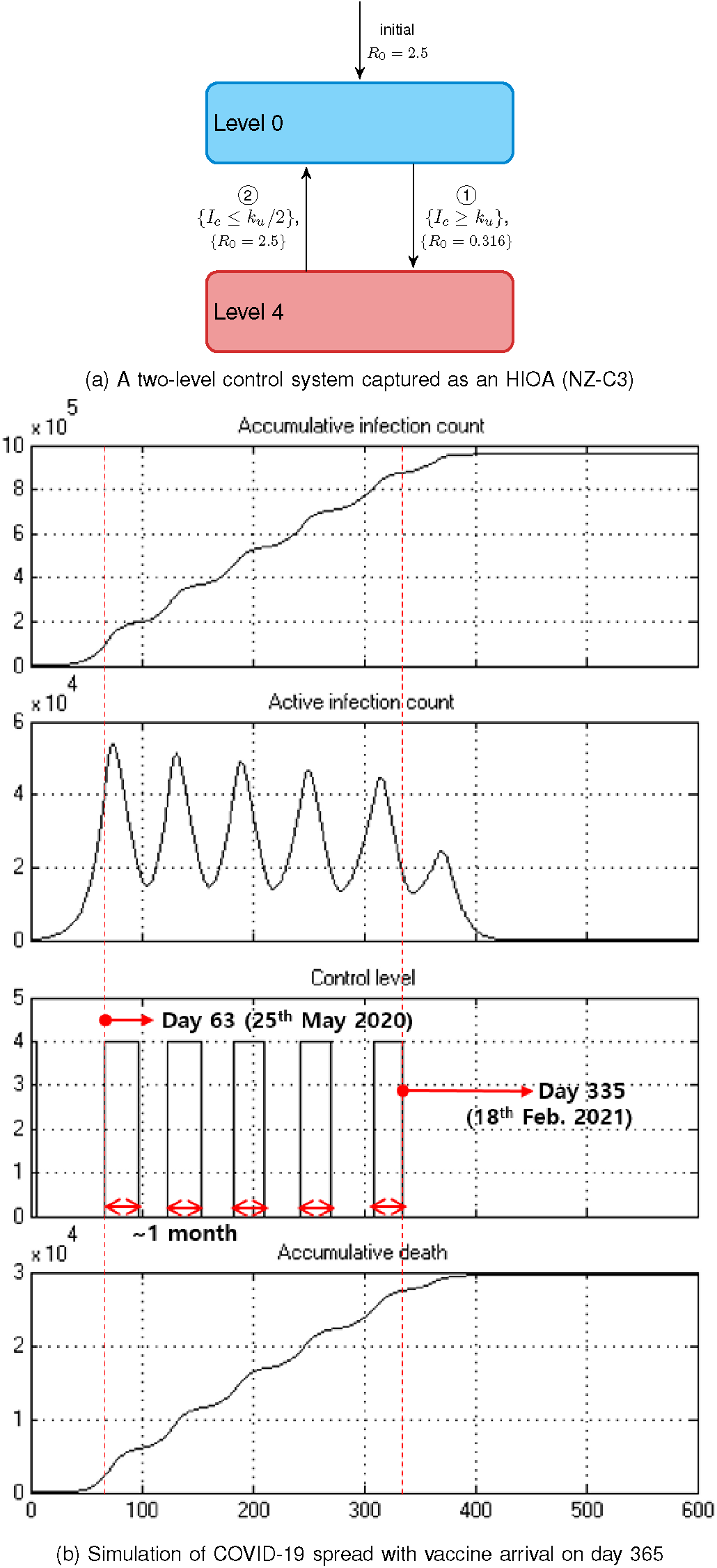
Examples of government interventions with only lockdown action

The simulation results for this model in our framework are shown in Figure 4b. As expected, the control level frequently switches between four and zero as the number of active cases oscillates. Interestingly, a number of one-month lockdown happens after day 63. Although the peaks of oscillation in the infection case graph gradually diminishes over time, the final number of deaths is extremely high and control remains in the lockdown for a long period of time, causing drastic impacts on the economy.

## Discussion

The compositional approach of CCPE allows the flexibility of formal modelling and validation of government control strategies to manage a pandemic. We have shown the ability of CCPE to model the dynamics of COVID-19 in conjunction with the various intervention techniques that governments employ. Table I compares the controllers used in this paper. As we can see, in the case of New Zealand, the controller `NZ-C2` achieves much better overall outcome compared to the simple controller `NZ-C1`. While the lockdown for `NZ-C1` lasts for over 120 days to achieve a near-zero infection count, the economic impact of such a long lockdown may be catastrophic. In contract, the controller `NZ-C2` has a gradual lifting of restrictions, which reaches level 1 much faster. Also, the overall risk of this strategy is a marginal increase in the number of deaths. In contrast to these two controllers, is the third control strategy `NZ-C3`, which introduces oscillations. We can see immediately the impact of a poorly managed control strategy, which may lead to three orders of magnitude more deaths. Finally, we also present the controller for Italy, which is modelled based on the actions of their government and as reported in [13].

**TABLE I:** A summary of Compositional Cyber-Physical Epidemiology case studies

| Figure | Plant | Controller | Confirmed cases | Deaths | Description | Social Impact |
| --- | --- | --- | --- | --- | --- | --- |
| 1 | PL-1 | NZ-C1 | 1670 | 33 | Indefinite lockdown | Lockdown lasts for 120 days |
| 2 | PL-2 | NZ-C2 | 2459 | 49 | Four level control | Business can operate after day 39, and a near zero infection count is achieved on day 398 |
| 4 | PL-1 | NZ-C3 | 960,000 | 29,600 | Two level control | Infection count oscillates until it reaches zero on day 409 |
| 3 | PL-1 | I-C1 | 273,000 | 35,000 | Three level control | A near zero infection count is achieved after 420 days |

CCPE allows the formal modelling of complex controllers. This enables the systematic evaluation of various control strategies in order to determine the best approach for a country which minimises the economic and social impacts, in addition to achieving the best healthcare outcome.

While the CCPE framework as presented here is based on the SEIR model [12], there is nothing that restricts our framework to such a model. Any continuous model which can be captured through a series of ODEs is able to be used which can open the door to more accurate simulation techniques, such as the enhanced version used by CovidSIM [3], the recent SIDARTHE model [26], or even microscale modelling. We have already shown this ability by suggesting some modification to SEIR to better account for contact tracing and isolation. This is presented in the *Methods* section and is denoted as the revised plant model `PL-2` in Table I.

The effectiveness of the CCPE framework relies on the fidelity of the transmission model and parameter estimation, requiring expertise in both epidemiology and statistical analysis. As such, the estimation of *R*_0_ is technically challenging [27] and the value varies due to different model assumptions and estimation procedures [25], [28], [29], [30], [31]. While the World Health Organization (WHO) estimates that the basic *R*_0_ ranges between 1.4 to 2.5 [32], Liu et al. suggested that the value is expected to be higher based on evolving research [33].

To apply the CCPE framework to other countries, the *R*_0_ value should be examined. However, this reproduction number varies based on the control measures implemented by each country [27], [34], [28], [35], [36]. To investigate the interaction between government interventions and disease transmission dynamics, action-specific *R*_0_ values are essential. Apart from the control actions, many factors, such as population density [37], mobility [28], and spatial heterogeneity [38], affect the *R*_0_ value.

The ability for our CCPE framework to work across a range of these different country-specific plant models and various control designs creates a useful tool for designing strategies to fight COVID-19. The analysis of counter-measures and their impact on dealing with the disease has traditionally been limited to simple “if-else” style controllers, and here we show the ability to model counter-measures which are able to include some form of state in their logic.

## Outlook

In this work, we evaluated the composition of a controller with an epidemiological model. However, the CCPE framework is far more flexible. HIOA-based modelling can be composed with any number of other HIOA. Further HIOAs could be used which take into account aspects such as legislation, culture, economy structure, administration, etc. [39], [40], [41]. For example, an economic model could be added [2], which takes into account the various measures being applied in order to provide a metric of the financial toll. Such a model could then be used to design a controller which not just minimises the number of deaths in the population, but also reduces the economic impact in a form of bi-criteria optimisation [42], [43].

In our work, the criteria used for switching between modes of the controller were based on comparing the number of active cases to the ICU capacity. Instead, control mechanisms could be created which take into account additional information, such as the climate, to more accurately capture the decision-making process. Moreover, we could further refine the dynamical modes to better represent the rate of testing.

Finally, a robust estimation approach of action specific *R*_0_ values within context of geographical and social heterogeneity should be systematically investigated in the future. COVID-19 is still relatively new and there exists a large variation in potential reproduction numbers between studies. We note that the accuracy of any epidemiological model depends on the accuracy of its reproduction number, and so further improvements in this area would be of great benefit. For example, there is the potential for the adoption of an approach as recently proposed in [11] if the reproduction number could be approximated as a continuous function. While this is a challenging proposition, our work opens the door for more engineering researchers to create an impact on current and future pandemics. A momentum is already in evidence as reported in [44] to show how Engineers are coming together to contribute to this cause in various ways.

## Methods

### The SEIR model of COVID-19

The modified SEIR model [12] consists of variables which represent the various sub-populations during an epidemic: susceptible (*S*), exposed (*E*), pre-symptomatic (*P*), infectious (*I*), recovered (*R*), and deaths (*D*). The infectious and recovered cases are further categorized into untested (*I_u_*, *R_u_*) and confirmed (*I_c_*, *R_c_*). The dynamics of the transmission between these sub-populations can be described by a series of coupled ODEs, shown in Equations 1 through 8.

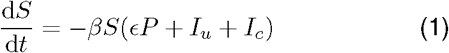

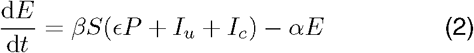

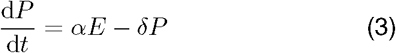

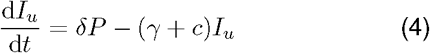

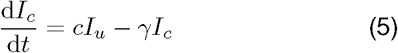

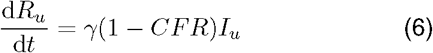

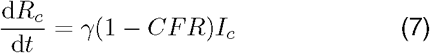

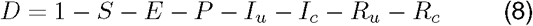

The Case Fatality Ratio (CFR) depends on the number of active people in the ICU and the ICU capacity. If the number of active people in the ICU is within the ICU capacity then the CFR is simply equal to some lower bound *CFR*_0_ (1 %). When this limit is exceeded, the CFR is decided by a mixture of patients who are receiving ICU care (*CFR*_0_) and those who are not (*CFR*_1_). The result of this is a piecewise function as in Equation 9 where *CFR*_1_ (2 %) is the maximum fatality rate, *n_ICU_* denotes the maximum ICU beds, *N* is the population size, and *p_ICU_* is the proportion of total cases which require ICU attention.

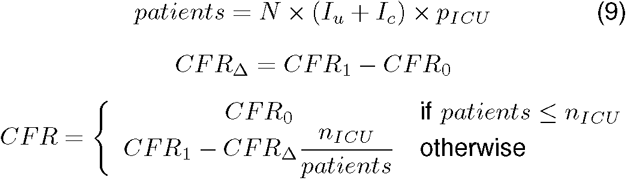

The SEIR model can be described as an HIOA, shown earlier in Figure 1b. *X* is the vector of all epidemic variables initialized to *X*_0_, A is the matrix of the parameters, and *Ẋ* = *AX* is the matrix representation of Equations 1 through 8). When the ICU demand *N_icu_* is less than or equal to the maximum ICU capacity *n_ICU_*, the HIOA stays in the location *Below ICU* with a CFR of *CFR*_1_. Otherwise, control goes to the location *Beyond ICU* and the CFR is defined by Equation 9.

In the model (Equations 1 and 2), the reproduction number *R*_0_ determines the transmission rate *β* as per Equation 10. Here, *∊* is the relative infectiousness in the presymptomatic period, *δ* is the transition rate from presymptomatic to infectious, and *γ* is the transition rate from infectious to recovered.

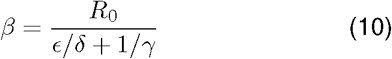

These transition rates are decided by the virus nature, while *R*_0_ depends on the contacts and the transmissibility [10]. The government control measures can impact this reproduction number, and hence also *β*, by reducing:

- physical contacts (e.g. travel restriction, self-isolation, work at home, close schools, etc.), or
- the transmissibility (e.g. hand washing, public disinfection efforts, etc.)

In order to start the propagation of the disease through the population we start with an initial number of cases (*I_c_*) which matches with the initial number of reported cases. Typically, our simulations start after a country has reached 100 total cases as this is a likely point where local transmission, if not community transmission, has started to occur. Additionally, this allows us to isolate the population from the rest of the world and ignore the potential inflow and outflow of infected people as travel is heavily restricted by this point in time.

We propose a revision of the recent SEIR model [12] in this paper to account for better management of the pandemic using improved case isolation and contact tracing. In Table I, we denote the SEIR model [12] as the plant model `PL-1` while our revised model is marked as the plant model `PL-2`. This is since case isolation and contact tracing could significantly reduce *R*_0_ for identified cases (i.e. *I_c_*) [35]. We use different parameters for the transmission rate *β* (Equations 1 and 2) such that the confirmed cases have lower transmissivity due to the combined effects of isolation and contact tracing. The resultant refined model replaces Equations 1 and 2 with Equations 11 and 12.

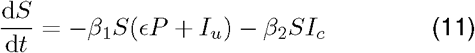

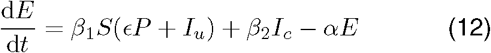

### The New Zealand model of COVID-19

For the epidemiological model of New Zealand, we use base reproduction number of 2.5 as is widely reported without control measures in place [25]. To investigate the interaction between government interventions and disease transmission dynamics, we need to introduce various reproduction numbers for the different action control strategies and stages. The estimation of *R*_0_ is technically challenging [27] and a number of studies have been done [45], [30], [33], [29], [36]. However, these values are not specific to certain control policies.

We identified which interventions are applied in the New Zealand alert levels, indicated in Table II by a tick (✓) or a cross (✗) to capture if a given intervention is applied (respectively not applied) in a given alert level. Each intervention is also weighted in its effectiveness, with the weighted sum being 2.184. A triangle (Δ) is used when an intervention is partially applied. In this case, half the weight is considered. At the bottom of Table II, we show the calculated reproduction numbers for each alert level by taking into account both the base reproduction number *R*_0_ and the interventions applied. In summary, the *R*_0_ values for alert levels 4 through 1 are 0.316, 0.827, 1.384, 1.570 respectively. The maximum value of *R*_0_ is 2.5, which corresponds to level 0.

**TABLE II:** A list of the interventions involved at each alert level in New Zealand, and the reproduction number derivation

| Intervention | Weight | Level 4 | Level 3 | Level 2 | Level 1 | Level 0 |
| --- | --- | --- | --- | --- | --- | --- |
| Widespread testing | 0.186 | ✓ | ✓ | ✓ | ✓ | ✗ |
| Temperature checkpoints | 0.093 | ✓ | ✓ | ✓ | ✓ | ✗ |
| Contact tracing | 0.186 | ✓ | ✓ | ✓ | ✓ | ✗ |
| Close contacts of confirmed cases ordered to self-isolate | 0.093 | ✓ | ✓ | ✓ | ✓ | ✗ |
| Large scale disinfection efforts | 0.046 | ✓ | ✓ | ✓ | ✗ | ✗ |
| Distribution of PPE to at-risk workers | 0.093 | ✓ | ✓ | ✓ | ✓ | ✗ |
| Hygiene public awareness efforts | 0.186 | ✓ | ✓ | ✓ | ✓ | ✗ |
| International travel ban | 0.186 | ✓ | ✓ | △ | ✗ | ✗ |
| Domestic travel restrictions | 0.093 | ✓ | ✓ | △ | ✗ | ✗ |
| People forced to remain home | 0.186 | ✓ | ✗ | ✗ | ✗ | ✗ |
| Bans on outdoor gatherings over 500 people | 0.093 | ✓ | ✓ | ✓ | ✓ | ✗ |
| Bans on indoor gatherings over 100 people | 0.093 | ✓ | ✓ | ✗ | ✗ | ✗ |
| Bans on recreational sports | 0.046 | ✓ | ✓ | ✗ | ✗ | ✗ |
| Bars and restaurants close | 0.186 | ✓ | △ | ✗ | ✗ | ✗ |
| Schools close | 0.186 | ✓ | △ | ✗ | ✗ | ✗ |
| Tertiary education facilities close | 0.093 | ✓ | △ | ✗ | ✗ | ✗ |
| Small food retailers close | 0.093 | ✓ | ✗ | ✗ | ✗ | ✗ |
| Non-essential retail business close | 0.093 | ✓ | △ | ✗ | ✗ | ✗ |
| Summation | 2.184 | 2.184 | 1.673 | 1.116 | 0.930 | 0 |
| Base reproduction number ($R_0$) | | 2.5 | 2.5 | 2.5 | 2.5 | 2.5 |
| Final R value |  | 0.316 | 0.827 | 1.384 | 1.570 | 2.5 |

The controller `NZ-C2`, in Figure 2a, matches a given alert level to its corresponding *R*_0_ value. Initially, the control starts from `Pre-LD` and move to `LD` just like the previous controller in Figure 1c. After 33 days this corresponds to 27th April 2020, which is the scheduled start of `Level 3`. After this point, the control enters `Level 3`, and the reproduction number is set to 0.827. The transitions from `Pre-LD` to `LD` and `LD` to `Level 3` are taken based on time, like `NZ-C1` since these mimic known time based government actions.

Subsequently, the government decisions, which are yet unknown, will have to be mimicked using more complex mechanisms. We use the following strategy to determine the transition conditions, which will not be time based alone, as follows. First we denote the rate of new cases per day as *Ċ*, the current number of infected cases is *I_c_*, and *k_u_* denote the upper bound values of hospital capacity. We consider the following parameter values based on the published data from New Zealand [12]. We set *k_l_*_3_, *k_l_*_2_, and *k_l_*_1_ as 500, 250, and 50, respectively. The maximum hospital capacity *k_u_* is 40,000. *dk_l_*_3_, *dk_l_*_2_, and *dk_l_*_1_ are 10, 5, and 0.01, respectively. Finally, in our results, we assume that a vaccine will arrive 365 days after 20th March 2020. At this time, the number of susceptible people decreases to zero, assuming widespread adoption of an effective vaccine.

The conditions for increasing the alert level are based on the current number of infected cases (*I_c_*). For example, from level two if *I_c_* ≥ *k_l_*_3_ then the alert level immediately rises to three. On the other hand, the alert level can go down if the increasing rate of new cases per day (*Ċ*) is less than a certain amount. For example, from level three if *Ċ* ≤ *dk_l_*_3_ then the alert level decreases to level two. In addition, to avoid frequent oscillations between levels, a minimum duration within a level before being able to drop down to a lower level is added and is set to be either 15 or 30 days.

From `Level 3`, the alert level can go down to `Level 2` if the increasing rate of new cases per day (*Ċ*) is less than a control parameter *dk_l_*_3_. For transition ③, 30 days is the minimum time of remaining in `Level 3` before entering `Level 2`. This timing constraint is included to avoid undesirable switching between levels. On this transition, the reproduction number is set to 1.384.

### The Italy model of COVID-19

Unlike New Zealand, Italy does not issue a systemic intervention strategy for COVID-19. Instead, the government releases the actions incrementally as they are needed. The Oxford COVID-19 Government Response Tracker (OxCGRT) [13] provides a stringency index of the measures taken by various governments around the world. According to the stringency index of Italy’s interventions, we divide the transmission trajectory into four phases. Considering that the initially reported cases are mostly imported rather than community transmission, the starting point of the analysis is 23rd February 2020, when the reported number of cases is 155. As a first attempt, we use the SEIR model [12] and curve fitting to estimate policy-specific reproduction numbers for Italy. We use the MATLAB^®^ function `lsqcurvefit` to search for these reproduction numbers for each phase by minimizing the square of the residual error between the SEIR simulation and the reported data [46]. The resulting reproduction numbers are listed in Table III.

**TABLE III:** Estimated reproduction numbers for each phase in Italy

| Phase 0 | Phase 1 | Phase 2 | Phase 3 |
| --- | --- | --- | --- |
| 23 Feb to 4 Mar | 4 Mar to 10 Mar | 10 Mar to 20 Mar | 20 Mar to 7 Apr |
| $R_{L0}$ | $R_{L1}$ | $R_{L2}$ | $R_{L3}$ |
| 6.3533 | 4.8051 | 3.2693 | 0.5216 |

The controller is shown in Figure 3a. For dropping alert levels, we have values of 10, 5, and 0.01 for *dk_l_*_3_, *dk_l_*_2_, and *dk_l_*_1_ respectively. The population of Italy (*N*) is 60,461,828 and we have level changing constraints of 6046, 3023, and 605 for *k_l_*_3_, *k_l_*_2_, and *k_l_*_1_ respectively. Additionally, the constraint to level four (*k_u_*) is equal to the hospital capacity of approximately 483,694 [12].

### HAML

Hybrid Automata Modelling Language (HAML) [21] is a recently developed tool in our group for the compositional modelling and verification of CPSs. To create the CCPE system in HAML we simply create two automata, one each for the plant and controller, and compose them as a single network. For the plant model, Listing 1a, we have an automata with an input *R*_0_ value which is used to determine the rate of reproduction in the model. Additionally, there are two outputs for the number of currently infected (and tested) people, *I_c_*, and the rate of change in the number of cases (*C_dot_*). The two locations of Figure 1b are shown which have the same flow constraints but differ in their calculation of the CFR to create a piecewise implementation of Equation 9 through the use of update constraints.

The discrete controller has external inputs and outputs which mirror those of the plant model, having two inputs, *I_c_* and *C_dot_*, and a single output, *R_0_*. Listing 1b shows this controller captured in HAML, using locations for each of discrete modes that it can be in. When the number of current confirmed cases (*I_c_*) reaches an upper bound for each location then the control progresses to a higher alert level, while when the change in number of cases (*C_dot_*) reaches a lower bound then control transitions to a lower alert level. The values of *R*_0_ for each control location are taken from Table II.

Finally, composition between these two components simply requires mapping their respective inputs and outputs together. This is achieved by defining each of the previous models, creating a single instance for each, and then providing the mapping of their variables, as shown in Listing 1c.

**Listing 1:**
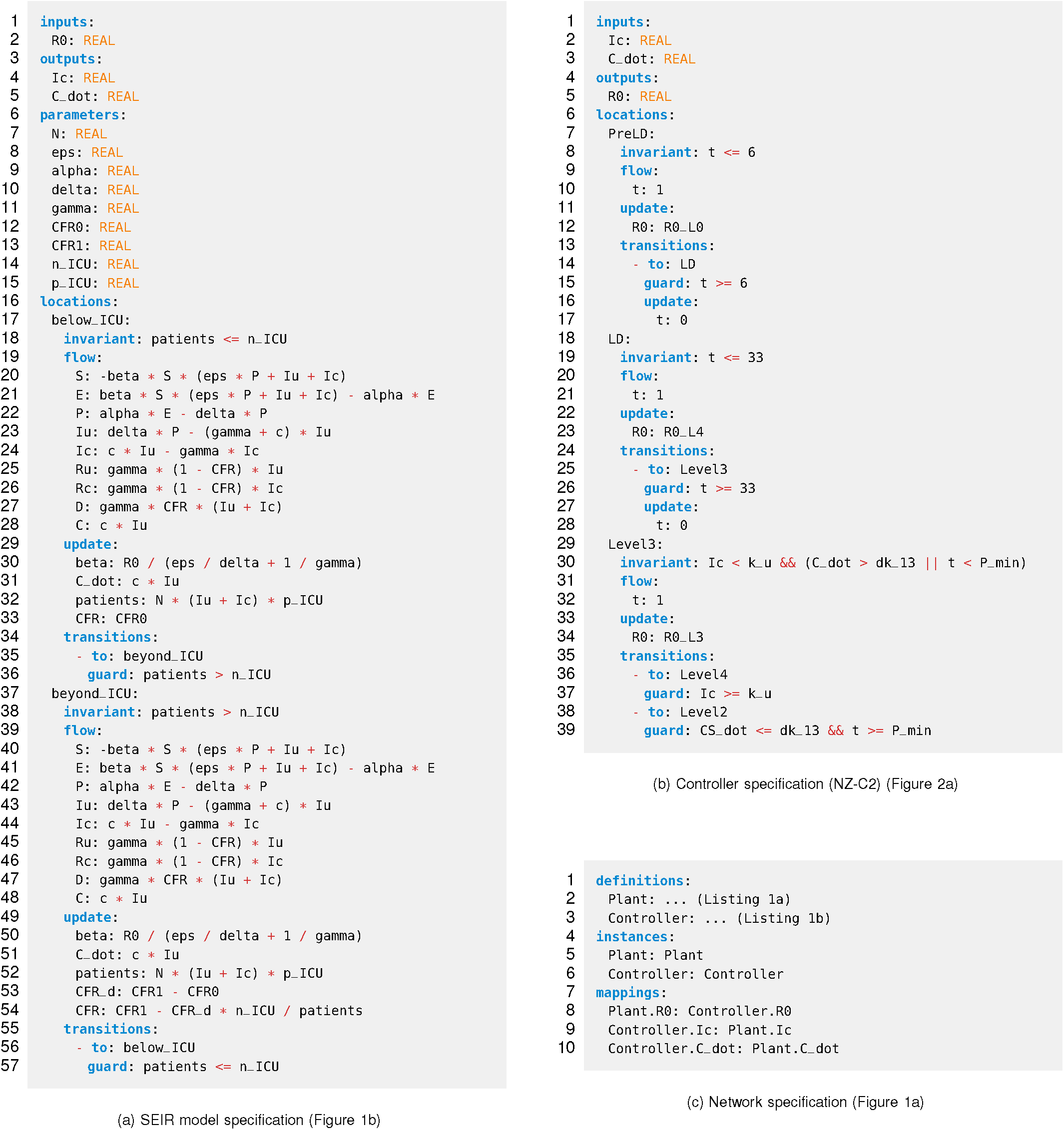
Example HAML specifications for the Compositional Cyber-Physical Epidemiology system

## Data Availability

The models used for this work are publicly available.

https://github.com/PRETgroup/ccpe-covid19

